# CLINICAL PROFILES OF GLAUCOMATOUS PATIENTS WITH HIGH- AND LOW-TENSION OPTIC DISC HEMORRHAGES: A COMPARATIVE STUDY

**DOI:** 10.1101/2020.09.05.20188904

**Authors:** Izabela Almeida, Diego Torres Dias, Paula Azevedo Alhadeff, Flavio Siqueira Santos Lopes, Carolina P B Gracitelli, Augusto Paranhos, Robert Ritch, Tiago Santos Prata

## Abstract

**Background/Aims:** Optic disc hemorrhage (DH) is an important glaucoma risk factor, and occurs in a wide intraocular pressure (IOP) range. We sought to characterize distinct clinical subtypes of patients with high- (HTDH) and low-tension DHs (LTDH).

**Methods:** In this cross-sectional study, treated glaucomatous patients with DHs from two Glaucoma Services were consecutively enrolled. Disc photographs were evaluated for the presence of DH by two glaucoma specialists. After inclusion, patients were classified on HTDH (IOP ≥ 16mmHg) and LTDH (IOP< 16mmHg; median split). Clinical and ocular data from the time of DH detection were compared between groups.

**Results:** One hundred thirty-three DH patients were included (LTDH = 66 eyes; HTDH = 67 eyes). Patients with LTDH were more often women than those with HTDH (77% vs 42%; p = 0.030). There was also a trend for a higher prevalence of Asian descendants (24% vs 9%; p = 0.058) and symptoms suggestive of vascular dysregulation (34% vs 14%; p = 0.057) in LTDH patients. Eyes with LTDH also had worse visual field (VF) mean deviation index (p = 0.037), higher prevalence of normal-tension glaucoma (NTG) diagnosis (46% vs 17%; p< 0.001) and tended to have thinner central corneas (p = 0.066).

**Conclusion:** Patients developing DHs with treated IOPs in the low teens seem to more frequently fit in a profile represented by women, NTG diagnosis and greater VF loss. The presence of symptoms suggestive of vascular dysregulation and race also seem to differ between these two clinical subtypes. A closer optic disc surveillance is recommended for patients with the LTDH subtype, as they may develop DHs despite seemingly well-controlled IOP.

## BACKGROUND

Glaucoma is a progressive chronic optic neuropathy and one of the leading causes of irreversible blindness worldwide. The diagnosis is usually established based on functional and structural damage, documented through visual field (VF) testing, and by optic nerve head (ONH) and retinal nerve fiber layer (RNFL) evaluation, respectively.^1^

Identification of ocular and systemic risk factors associated with glaucoma development and progression has been the focus of numerous studies.^2–8^ In this context, elevated intraocular pressure (IOP) remains the most important known risk factor and the only modifiable one. Currently, it is believed that IOP plays an important role in glaucoma development and progression across the entire glaucoma spectrum, even when IOP is statistically normal.^9–12^ Some of the previously identified risk factors seem to differ between eyes with elevated IOP and those with IOP within the normal range, though. For instance, older age, thinner corneas, black race and worse VF damage at baseline have been reported as risk factors for primary open-angle glaucoma development and progression,^3–5,7,8^ while migraine and female gender have been reported as normal-tension glaucoma (NTG) risk factors.^2^ Conversely, optic disc hemorrhage (DH) is one ocular risk factor that is significantly associated with disease development and progression independent of the glaucoma subtype.^4,5,13–17^

Since the first report by Bjerrum more than 100 years ago,^18,19^ the association between DH and glaucomatous damage has been the subject of numerous publications.^2,4,13–15,17–22^ Although its underlying causative mechanisms are still not well established, previous reports suggest that DHs might be associated with both mechanical and vascular phenomena. Characterized by splinterlike or flame-shaped hemorrhages at the border of the ONH, DH are rarely found in normal eyes and are observed more commonly in patients with NTG.^20–22^ However, DHs can occur over a wide range of IOPs.^4,13–15,17,20–22^

We hypothesized that the clinical profile of a patient who develops a DH whose IOP is in the low teens (low-tension disc hemorrhage [LTDH]) could be different from the profile of a patient that develops a DH with higher IOP values (high-tension disc hemorrhage [HTDH]). Identification of the clinical characteristics of these two different groups could help clinicians in the surveillance of such patients. Therefore, in the present study, we sought to characterize two distinct clinical profiles (LTDH and HTDH) of glaucomatous patients based on the IOP presented at the time of DH detection.

## METHODS

This protocol adhered to the tenets of the declaration of Helsinki and was approved by the ethics committee and the institutional review board of the New York Eye and Ear Infirmary and the Federal University of São Paulo.

### Participants

In this cross-sectional comparative study, we enrolled consecutive glaucomatous patients with DH from the Glaucoma Services of the two above-mentioned institutions. First, disc photographs of all patients were reviewed by 2 glaucoma specialists for the presence of DH. In cases of disagreement, the impression of a third investigator was used for adjudication. A DH was defined as a splinterlike or flame-shaped hemorrhage on or within the RNFL or neuroretinal rim. If peripheral to the disc margin, the hemorrhage needed to be contiguous to the β-zone parapapillary atrophy (PPA) when this feature was present.^23,24^ Exclusion criteria were the presence of diabetic retinopathy, vascular occlusive disease, optic disc drusen, ocular trauma and recent history of posterior vitreous detachment.

All participants had undergone a comprehensive ophthalmological evaluation, including best-corrected visual acuity, slit-lamp biomicroscopy, IOP measurement, gonioscopy, dilated fundoscopy, VF testing (24-2 Swedish interactive threshold algorithm, Humphrey Field Analyzer II; Carl Zeiss Meditec, Inc., Dublin, CA), optic disc stereophotographs, and color/red-free fundus imaging. Glaucomatous optic neuropathy was defined as cup-to-disc ratio>0.6, asymmetry between eyes ≥0.2, presence of localized defects of the RNFL, and/ or neuroretinal rim in the absence of any other anomalies that could explain such findings. Characteristic glaucomatous VF defect was defined as glaucoma hemifield test results outside normal limits or presence of at least 3 contiguous test points within the same hemifield on the pattern deviation plot at p< 1%, with at least 1 at p< 0.5%, excluding points on the edge of the field or those directly above and below the blind spot (based on two reliable VF tests).^24^

Patients were then divided into LTDH and HTDH groups according to the median IOP of the study population (16 mmHg). Patients presenting with an IOP ≥16 mmHg at the time of DH detection were classified as HTDH and those with an IOP< 16 mmHg were classified as LTDH. This methodology (median split) has been previously adopted in the literature for assessment of risk factors for glaucoma.^4^ Finally, clinical and ocular data from the time of DH detection were compared between patients with LTDH and HTDH. Whenever both eyes were eligible, one was randomly selected for analysis.

### Data collection, visual field and optic disc assessment

The baseline time point for purposes of data acquisition was defined as the date at which the initial DH was documented photographically. Baseline data included age, race, gender, IOP, corneal central thickness (CCT), refractive error, VF mean deviation index (MD), presence of parafoveal scotoma (PFS), optic disc phenotype, presence of PPA and RNFL defects, type of glaucoma, DH location and DH recurrence. Patient-related history of systemic vascular diseases (systemic hypertension or diabetes) and sings/symptoms of primary vascular dysregulation^25,26^ (PVD; migraine, Raynaud’s phenomenon, cold hands and low blood pressure) were also documented.

The classifications of optic disc phenotype^27^ and the presence of PFS^28^ were based on previously published studies. Briefly, discs were classified into 4 subtypes: focal loss (type 1), myopic (type 2), senile sclerotic (type 3) and generalized cup enlargement (type 4).^27^ PFS was defined as a VF defect in one hemifield within 10^0^ of fixation with at least one point at p< 1% lying at the two innermost paracentral points, with or without defects outside the central 10^0^ in the superior or inferior arcuate area.^24,28^

### Statistical analysis

Descriptive analysis was used to present demographic and clinical data. D’Agostino-Pearson’s test was performed to determine whether data had a normal distribution. Descriptive statistics included mean and standard deviation for normally distributed variables and median, quartiles for those non-normally distributed. Independent samples *t* test was used to compare continuous normally distributed variables between groups, while the Mann-Whitney test was used to compare those non-normally distributed. Categorical data were compared using χ^2^ test. Computerized analysis was performed using MedCalc software (MedCalc Inc., Mariakerke, Belgium). The alpha level (type I error) was set at 0.05.

## RESULTS

One hundred thirty-three glaucomatous patients (133 eyes) with DH were included in the analyses. The division according to IOP values at DH detection resulted in 66 eyes with LTDH and 67 eyes with HTDH (table 1). The LTDH group had a significantly higher frequency of women (77% vs 42%; p = 0.030), NTG patients (46% vs 17%; p< 0.001) and worse MD values (−5.2 vs −3.3 dB; p = 0.037) when compared to the HTDH group. There was also a trend for a higher prevalence of Asian descendants (24% vs 9%; p = 0.058), thinner CCT (518 vs 531 microns; p = 0.066) and symptoms suggestive of PVD (34% vs 14%; p = 0.057) in LTDH patients compared to those with HTDH.

**Table 1.** Characteristics of Glaucomatous Patients with Low-Tension and High-tension Disc Hemorrhages.

|  | LTDH (n=66) | HTDH (n=67) | P value |
| --- | --- | --- | --- |
| Age (years) | 67.5±9.7 | 65.6±12.4 | 0.326 |
| Gender (M/W) | 23%/77% | 58%/42% | 0.030 |
| Race (Wh/AD/A/Others) | 66%/6%/24%/4% | 69%/9%/9%/13% | 0.058 |
| Presence of PFS | 63% | 51% | 0.308 |
| Visual Field Mean Deviation (dB) | -5.2 (-12.4; -2.5) | -3.3 (-5.8; -1.8) | 0.037 |
| DH Recurrence | 25% | 19% | 0.574 |
| DH location (IT/ST/IN/SN) | 79%/15%/0%/6% | 81%/15%/4%/0% | 0.845 |
| Presence of PPA | 84% | 83% | 0.831 |
| Localized RNFL Defect | 67% | 74% | 0.776 |
| Central Corneal Thickness (μm) | 518 (501 – 541) | 531 (501 – 569) | 0.066 |
| Intraocular Pressure (mmHg) | 14 (12.5 – 14.5) | 19 (17 – 21) | <0.001 |
| Glaucoma Diagnoses (POAG/PACG/NTG/Others) | 29%/14%/46%/11% | 55%/6%/17%/22% | <0.001 |
| Optic Disc Phenotype (types 1/2/3/4)* | 42%/10%/15%/33% | 52%/5%/14%/29% | 0.309 |
| Systemic Vascular Diseases (DM and/or Hypertension) | 66% | 56% | 0.686 |
| PVD Symptoms | 34% | 14% | 0.057 |
| Spherical Equivalent (Diopters) | -0.5 (-1.7 – 1.0) | -0.0 (-1.25 – 1.0) | 0.458 |

Normally distributed data are provided as mean ± standard deviation and non-normally distributed data are provided as median and interquartile intervals whenever indicated.
LTDH= low-tension disc hemorrhage; HTDH= high-tension disc hemorrhage; M= men; W= women; Wh= White; AD= African Descendant; A= Asian; PFS= parafoveal scotoma; dB= decibels; DH = disc hemorrhage; IT = inferior-temporal; ST = superior-temporal; IN= inferior-nasal; SN= superior-nasal; PPA= parapapillary atrophy; RNFL = retinal nerve fiber layer; POAG= primary open-angle glaucoma; PACG= primary angle-closure glaucoma;
NTG= normal-tension glaucoma; DM= diabetes mellitus; PVD= primary vascular dysregulation.
\*Type 1 = focal loss; type 2 = myopic; type 3 = senile sclerotic; type 4 = generalized cup enlargement.

The most common optic disc phenotype was the focal loss subtype and the most frequent DH location was the inferior temporal sector in both groups. Nonetheless, these parameters did not significantly differ between LTDH and HTDH groups (p ≥0.309). Additionally, no significant differences were found regarding age, presence of PFS, systemic vascular diseases, presence of localized RNFL defects, β zone PPA, spherical equivalent and DH recurrences (p ≥0.308).

## DISCUSSION

Although many studies have analyzed the relationship between glaucoma and DHs, a detailed comparison between the clinical profiles of eyes with LTDH and HTDH has not been done yet. It is know that DHs are rarely found in normal eyes and, although they are more commonly observed in NTG patients, DHs may appear in a wide range of IOPs.^4, 13–15, 17, 20–22^ In this context, it would be reasonable to ask: do all these DHs represent the same clinical situation? Does a patient that develops a DH with an IOP as high as 25 mmHg have the same clinical characteristics as someone whose IOP was as low as 12 mmHg at the time of DH detection? In the present study, analyzing a large series of more than 130 patients with DH with different IOP levels, we sought to answer this question. Our data suggests that it is reasonable to characterize DH eyes into two distinct clinical subtypes, LTDH and HTDH, which seem to differ not only in some systemic characteristics (gender, race and symptoms of PVD), but also in ocular features (disease stage, NTG diagnosis and CCT). As far as we know, this is the first study to report such findings.

There are scant data in the literature regarding the comparison of clinical features between eyes developing DHs with low and high IOP values. At a first sight, this fact would preclude a straight comparison between our study findings and previously published reports. Nonetheless, a more careful analysis reveals that our results seem to corroborate and add to some findings previously documented in DH patients with lower IOP values. We will address these findings separately herein. For instance, Healey and colleagues,^21^ evaluating 3654 participants in a population-based study, observed that DH prevalence was higher in women than in men (OR, 1.9; CI, 1.0-3.5). In the same study, it was noted that the prevalence of DH in the NTG group was higher than in high-tension glaucoma patients (25% *versus* 8%, OR, 2.9; CI, 1.1-8.1). Although we have adopted a different methodology, in which patients were divided according to the IOP value documented at the time of DH detection, our findings also indicate a relationship between female gender and NTG diagnosis with DH, mainly in those developing DHs at low IOP values (eyes with LTDH).

Analyzing the relationship between the magnitude of glaucomatous damage and the presence of DH in the literature, we found some interesting reports. Although there are robust data indicating that DH occurrence is significantly related to disease development and progression,^13, 29, 36, 29^ only a few reports have investigated whether eyes with more advanced glaucoma would be more prone to develop a DH.^21,22,30^ Healey et al^21^ observed that DH were more frequent in eyes with larger vertical cup-disc ratios (OR, 2.2; CI, 1.1 - 4.6). Additionally, in the low-pressure glaucoma treatment study (LOTGS),^30^ a narrower neuroretinal rim width at baseline (HR, 2.91; P = .048) was an independent risk factor for DH detection. Overall, these studies suggest that eyes with more advanced disease (except for those without detectable neuroretinal rim)^22^ have a greater risk for DH occurrence. Our findings provide additional information, indicating that, in treated glaucomatous patients, those with greater VF loss (more advanced disease) are more likely to have a DH detected even at low IOPs (LTDH group).

Regarding vascular dysregulation, it has been consistently related to the glaucoma pathophysiology in the literature.^26,31,32^ Flammer and colleagues reviewed this subject and concluded that PVD might be not only an important factor for glaucoma development and progression, but also for a higher frequency of DH despite low IOP values.^26,31,32^ Additionally, in the LOTGS study, the presence of signs and symptoms of PVD (migraine, low mean systolic blood pressure and low mean arterial ocular perfusion pressure) were independent risk factors for DH detection.^30^ We believe our data corroborate these findings, as signs and symptoms of PVD were at least twice more frequent in eyes with LTDH.

In face of our results, we believe it is important to delve into the possible underlying causative mechanisms regarding DH occurrence. Although DH has been the subject of many studies, the exact mechanism by which they appear remains unclear. In this context, mechanical and vascular mechanisms have been proposed to explain DH occurrence. The mechanical theory proposes that DH results from the pressure gradient across the lamina cribrosa (LC) and high IOP-induced stress on the ONH. This would lead to mechanical shearing stress forces on the nerve fibers and capillaries at the level of LC pores, stretching of the anterior capillaries upon backward bowing of the LC and consequent bleeding.^11,33^ On the other hand, the vascular theory suggests that vascular dysregulation (either primary^34^ – PVD – or secondary – to diabetes mellitus, systemic arterial hypertension and platelet aggregation inhibition,^35^ for example) may be related with DH through local breakdown of the blood-retinal barrier due to basal membrane and endothelial cells dysfunction, microinfarctions in the optic disc head, and consequent bleeding.^32^ Increased levels of circulating endothelin-1 (a tissue hormone with vasoactive properties) and matrix metalloproteinases-9 (involved in tissue remodelling) may be related in the pathogenesis of both PVD and DH.^34^ Although both mechanical and vascular mechanisms may be present in eye DH eyes, our results support the idea that LTHD patients may represent a population in which the vascular mechanisms have a more prominent role than the mechanical mechanism. First, because of IOP itself, as these patients frequently had IOP in a previously considered “safe” range in the low-teens before the DH occurrence and, therefore, the IOP itself would be expected to not cause intense mechanical shearing forces at LC level. Second, because of the specific demographic characteristics we documented in the LTDH group (higher prevalence of women, Asian descendants, NTG diagnosis and PVD symptoms). In fact, these Asian, middle-aged women with NTG profile has been previously linked with PVD, along with other symptoms and signs, such as cold hands and feet, migraine and Raynaud’s phenomenon.^26,36,37^

At this point it is important to briefly discuss the main clinical implications of our findings. We believe they are directly related to the clinical management and monitoring of glaucomatous patients, based on the following rationale. It is well known that DH can occur in a wide range of IOP values, and have been consistently associated with glaucoma development and progression. Although DH usually indicates poor disease control, it may occur even in eyes with seemingly well-control IOP (low-teens).^2,14,15,17,38–43^ Therefore, the knowledge of the clinical profile of eyes that may present DH even at low IOP values, may help clinicians in the surveillance of DH detection in these eyes. In this context, special attention must be given to glaucomatous patients with a LTDH profile (Asian, women, diagnosis of NTG and signs of vascular dysregulation) and greater VF loss (worse MD values).

The present study has some specific limitations that should be addressed. First, baseline IOP was based on a single measurement. A 24-hour tension curve would have been ideal in order to detect possible IOP peaks. Second, due to the retrospective nature of our data, we can establish possible associations, but not cause-effect relationships. Third, the prevalence of systemic risk factors was investigated on the basis of self-reported assessment, which might have reliability and accuracy issues. Finally, even though we have adopted strict criteria to define a glaucomatous DH in our study, some patients with DHs related to other (non-glaucomatous) conditions might have been included. These facts should be considered while interpreting our findings.

In conclusion, glaucomatous patients that present DHs with treated IOPs in the low-teens (< 16 mmHg) frequently fit in a profile represented by women, NTG diagnosis and worse MD index values. Thinner CCT, Asian ancestry and symptoms and signs of vascular dysregulation were also marginally associated with LTHD. We believe that this clinical profile of LTHD patients may represent a population of eyes in which vascular mechanisms would have a more prominent role than mechanical causes in DH occurrence. Furthermore, our findings may help clinicians in the identification and surveillance of patients at a higher risk for DH – and consequent glaucoma progression – even with IOP in the low-teens and seemingly well controlled.

## Data Availability

All data referred to in the manuscript is available

## Reference

1. Weinreb RN, Khaw PT. Primary open-angle glaucoma. Lancet. 2004;363(9422): 1711-1720.

2. Drance S, Anderson DR, Schulzer M, Collaborative Normal-Tension Glaucoma Study G. Risk factors for progression of visual field abnormalities in normal-tension glaucoma. American journal of ophthalmology. 2001; 131(6): 699-708.

3. Gordon MO, Beiser JA, Brandt JD, et al. The Ocular Hypertension Treatment Study: baseline factors that predict the onset of primary open-angle glaucoma. Archives of ophthalmology. 2002; 120(6): 714-720; discussion 829-730.

4. Leske MC, Heijl A, Hyman L, et al. Predictors of long-term progression in the early manifest glaucoma trial. Ophthalmology. 2007; 114(11): 1965-1972.

5. Miglior S, Torri V, Zeyen T, et al. Intercurrent factors associated with the development of open-angle glaucoma in the European glaucoma prevention study. American journal of ophthalmology. 2007; 144(2): 266-275.

6. Musch DC, Gillespie BW, Lichter PR, Niziol LM, Janz NK, Investigators CS. Visual field progression in the Collaborative Initial Glaucoma Treatment Study the impact of treatment and other baseline factors. Ophthalmology. 2009; 116(2): 200-207.

7. Nakagami T, Yamazaki Y, Hayamizu F. Prognostic factors for progression of visual field damage in patients with normal-tension glaucoma. Japanese journal of ophthalmology. 2006; 50(1): 38-43.

8. Nouri-Mahdavi K, Hoffman D, Coleman AL, et al. Predictive factors for glaucomatous visual field progression in the Advanced Glaucoma Intervention Study. Ophthalmology. 2004; 111(9): 1627-1635.

9. Comparison of glaucomatous progression between untreated patients with normal-tension glaucoma and patients with therapeutically reduced intraocular pressures. Collaborative Normal-Tension Glaucoma Study Group. American journal of ophthalmology. 1998; 126(4): 487-497.

10. The effectiveness of intraocular pressure reduction in the treatment of normal-tension glaucoma. Collaborative Normal-Tension Glaucoma Study Group. American journal of ophthalmology. 1998; 126(4): 498-505.

11. Burgoyne CF, Downs JC, Bellezza AJ, Suh JK, Hart RT. The optic nerve head as a biomechanical structure: a new paradigm for understanding the role of IOP-related stress and strain in the pathophysiology of glaucomatous optic nerve head damage. Progress in retinal and eye research. 2005; 24(1): 39-73.

12. Kass MA, Heuer DK, Higginbotham EJ, et al. The Ocular Hypertension Treatment Study: a randomized trial determines that topical ocular hypotensive medication delays or prevents the onset of primary open-angle glaucoma. Archives of ophthalmology. 2002; 120(6): 701-713; discussion 829-730.

13. Budenz DL, Anderson DR, Feuer WJ, et al. Detection and prognostic significance of optic disc hemorrhages during the Ocular Hypertension Treatment Study. Ophthalmology. 2006; 113(12): 2137-2143.

14. Gordon J, Piltz-Seymour JR. The significance of optic disc hemorrhages in glaucoma. Journal of glaucoma. 1997; 6(1): 62-64.

15. Ishida K, Yamamoto T, Sugiyama K, Kitazawa Y. Disk hemorrhage is a significantly negative prognostic factor in normal-tension glaucoma. American journal of ophthalmology. 2000; 129(6): 707-714.

16. Leske MC, Heijl A, Hussein M, et al. Factors for glaucoma progression and the effect of treatment: the early manifest glaucoma trial. Archives of ophthalmology. 2003; 121(1): 48-56.

17. Siegner SW, Netland PA. Optic disc hemorrhages and progression of glaucoma. Ophthalmology. 1996; 103(7): 1014-1024.

18. Bjerrum J. Om en Tilfojelse til den Saedvanlige Synsfelfundersogelse samt om Synfeltet ved Glaukom. Nord Ophthalmol Tidsskr. 1889; 2: 141-185.

19. Bjerrum J. Om Glaukomets Kliniske Afgraensning. Nord Ophthalmol Tidsskr. 1892; 5: 129-138.

20. Drance SM, Fairclough M, Butler DM, Kottler MS. The importance of disc hemorrhage in the prognosis of chronic open angle glaucoma. Archives of ophthalmology. 1977; 95(2): 226-228.

21. Healey PR, Mitchell P, Smith W, Wang JJ. Optic disc hemorrhages in a population with and without signs of glaucoma. Ophthalmology. 1998; 105(2): 216-223.

22. Jonas JB, Xu L. Optic disk hemorrhages in glaucoma. Am J Ophthalmol. 1994; 118(1): 1-8.

23. Airaksinen PJ, Mustonen E, Alanko HI. Optic disc hemorrhages. Analysis of stereophotographs and clinical data of 112 patients. Archives of ophthalmology. 1981; 99(10): 1795-1801.

24. Dias DT, Almeida I, Sassaki AM, et al. Factors associated with the presence of parafoveal scotoma in glaucomatous eyes with optic disc hemorrhages. Eye (Lond). 2018; 32(10): 1669-1674.

25. Pache M, Flammer J. A sick eye in a sick body? Systemic findings in patients with primary open-angle glaucoma. Surv Ophthalmol. 2006; 51(3): 179-212.

26. Grieshaber MC, Mozaffarieh M, Flammer J. What is the link between vascular dysregulation and glaucoma? Surv Ophthalmol. 2007;52 Suppl 2: S144-154.

27. Broadway DC, Nicolela MT, Drance SM. Optic disk appearances in primary open-angle glaucoma. Survey of ophthalmology. 1999;43 Suppl 1: S223-243.

28. Park SC, De Moraes CG, Teng CC, Tello C, Liebmann JM, Ritch R. Initial parafoveal versus peripheral scotomas in glaucoma: risk factors and visual field characteristics. Ophthalmology. 2011; 118(9): 1782-1789.

29. Prata TS, De Moraes CG, Teng CC, Tello C, Ritch R, Liebmann JM. Factors affecting rates of visual field progression in glaucoma patients with optic disc hemorrhage. Ophthalmology. 2010; 117(1): 24-29.

30. Furlanetto RL, De Moraes CG, Teng CC, et al. Risk factors for optic disc hemorrhage in the low-pressure glaucoma treatment study. Am J Ophthalmol. 2014; 157(5): 945-952.

31. Flammer J, Haefliger IO, Orgul S, Resink T. Vascular dysregulation: a principal risk factor for glaucomatous damage? J Glaucoma. 1999; 8(3): 212-219.

32. Grieshaber MC, Terhorst T, Flammer J. The pathogenesis of optic disc splinter haemorrhages: a new hypothesis. Acta Ophthalmol Scand. 2006; 84(1): 62-68.

33. Quigley HA, Addicks EM, Green WR, Maumenee AE. Optic nerve damage in human glaucoma. II. The site of injury and susceptibility to damage. Archives of ophthalmology. 1981; 99(4): 635-649.

34. Flammer J, Konieczka K, Flammer AJ. The primary vascular dysregulation syndrome: implications for eye diseases. The EPMA journal. 2013; 4(1): 14.

35. Grodum K, Heijl A, Bengtsson B. Optic disc hemorrhages and generalized vascular disease. Journal of glaucoma. 2002; 11(3): 226-230.

36. Esporcatte BL, Tavares IM. Normal-tension glaucoma: an update. Arq Bras Oftalmol. 2016; 79(4): 270-276.

37. Mallick J, Devi L, Malik PK. Update on Normal Tension Glaucoma. J Ophthalmic Vis Res. 2016; 11(2): 204-208.

38. De Moraes CG, Prata TS, Liebmann CA, Tello C, Ritch R, Liebmann JM. Spatially consistent, localized visual field loss before and after disc hemorrhage. Investigative ophthalmology & visual science. 2009; 50(10): 4727-4733.

39. Diehl DL, Quigley HA, Miller NR, Sommer A, Burney EN. Prevalence and significance of optic disc hemorrhage in a longitudinal study of glaucoma. Archives of ophthalmology. 1990; 108(4): 545-550.

40. Kono Y, Sugiyama K, Ishida K, Yamamoto T, Kitazawa Y. Characteristics of visual field progression in patients with normal-tension glaucoma with optic disk hemorrhages. American journal of ophthalmology. 2003; 135(4): 499-503.

41. Rasker MT, van den Enden A, Bakker D, Hoyng PF. Deterioration of visual fields in patients with glaucoma with and without optic disc hemorrhages. Archives of ophthalmology. 1997; 115(10): 1257-1262.

42. Sung KR. Disc hemorrhage: is that a risk factor or sign of progression? Journal of glaucoma. 2012; 21(4): 275-276.

43. Xu L, Wang YX, Yang H, Jonas JB. Follow-up of glaucomatous eyes with optic disc haemorrhages: the Beijing Eye Study. Acta ophthalmologica. 2009; 87(2): 235.

